# Links between Cannabinoid Hyperemesis Syndrome symptoms and drug use, mental health problems, antisocial behavior, and personality in a national survey of adults in the United States

**DOI:** 10.64898/2026.02.26.26347188

**Authors:** Brian M. Hicks, Amanda M. Price, Blair J. Whittington, Paula Goldman, Mark A. Ilgen

**Affiliations:** Addiction Center, Department of Psychiatry, University of Michigan; Center for Clinical Management Research, Department of Veterans Affairs; Department of Pediatrics, University of Michigan

**Keywords:** cannabis, marijuana, nausea, vomiting, psychopathology, depression, personality

## Abstract

**Background:** Cannabinoid hyperemesis syndrome (CHS) is characterized by episodes of severe nausea, vomiting, and abdominal pain among those with heavy cannabis use. We estimated differences between those reporting CHS symptoms and those who use cannabis daily and less frequently on drug use, psychiatric problems, other health problems, antisocial behavior, and personality.

**Methods:** The National Firearms, Alcohol, Cannabis, and Suicide survey was administered to 7034 US adults (3427 male, 3607 female) in 2025. Survey items assessed substance use, common psychiatric symptoms, personality traits, and symptoms of CHS.

**Results:** Those with CHS symptoms reported the highest rates and greatest variety of drug use compared to others who used cannabis. Those with CHS symptoms reported higher rates of other drug use than those who used cannabis daily without CHS symptoms across a variety of drug classes, including opioids, hallucinogens, and sedatives, higher rates of drug overdoses, and greater use of all drug classes than those with less-than-daily cannabis use. Those with CHS symptoms also reported more depression, anxiety, sleep problems, chronic pain, antisocial behavior, intimate partner violence, and disinhibited personality traits than those who used daily (mean *d* = 0.58) and less frequently (mean *d* = 0.69) and those with no cannabis use in the past 12 months (mean *d* = 0.99).

**Conclusions:** Those with CHS symptoms exhibit a variety of psychological and behavioral problems including higher rates of other drug use, psychiatric symptoms, antisocial behavior, and dysfunctional personality traits. Results highlight the importance of understanding and addressing the broader psychosocial challenges faced by people experiencing CHS symptoms.

## 1. INTRODUCTION

Patterns of cannabis use have shifted dramatically in the United States over the past two decades with increasing access to new cannabinoid products, broader availability of high potency cannabis, and declining perceptions of risk.(ElSohly et al., 2016; Hall & Lynskey, 2020; Spindle et al., 2019) These changes have coincided with increases in the prevalence of cannabis use, with an especially striking increase in the prevalence of heavy cannabis use.(Caulkins, 2024; Mattingly et al., 2024; Wang et al., 2024) Cannabinoid hyperemesis syndrome (CHS) is one potential consequence that was first described in 2004 among those who use cannabis frequently over an extended time period.(Allen et al., 2004) CHS involves repeated nausea during the prodromal phase and recurrent episodes of severe nausea, vomiting, and abdominal pain during the acute phase.(Angulo, 2024; Hasler et al., 2025; Jiménez-Castillo et al., 2025; Loganathan et al., 2024; Sorensen et al., 2017; Stjepanović et al., 2025)

Research on CHS is still in a nascent state but this condition has been described with increasing frequency in acute care settings.(Garcia & Rodríguez, 2025; Jack et al., 2025) Using data from the National Firearms, Alcohol, Cannabis, and Suicide (NFACS) survey, we found that nearly 18% of those who reported a period of daily cannabis use in the past 5 years also endorsed episodes of severe nausea, vomiting, and abdominal pain suggestive of CHS, which translates to an estimated seven million US adults (Ilgen, 2026). This estimate indicates the population-level incidence of CHS symptoms may be much greater than previously understood based on the limited existing research from populations presenting to acute medical settings. Therefore, it is important to better characterize people with CHS symptoms in the general US adult population to improve an understanding of their unique needs and potential points of intervention.

What we know at present is that those with CHS symptoms tend to have lower income, lower educational attainment, younger age, and slightly more likely to be non-White race and female compared to other acute care patients and survey respondents.(Ilgen, 2026; Meltzer et al., 2025; Miki et al., 2025; Simonetto et al., 2012; Swartz & Franceschini, 2025) CHS symptoms are believed to be caused by heavy cannabis use. Consistent with this, a survey of people recruited from online support groups for CHS found that over 40% of people had used cannabis for five or more years prior to the onset of symptoms and a similar percentage (41%) reported using cannabis five or more times daily.(Meltzer et al., 2025) Our prior work also found that those with CHS symptoms reported more symptoms of cannabis use disorder even when compared to those with daily cannabis use without CHS symptoms.(Ilgen, 2026) More broadly, heavy cannabis use and cannabis use disorder have been associated with high rates of other drug use, alcohol use and problem use, tobacco smoking, antisocial behavior, depression, anxiety, and disinhibited and/or externalizing personality traits like impulsivity and aggression, along with poor sleep, chronic pain and poorer overall functioning.(Coelho et al., 2023; Copeland et al., 2022; Hasin, 2018; Hoch et al., 2025; Mannes et al., 2023; Solmi et al., 2023).

Based on these findings, we expected those reporting CHS symptoms to have high rates of other drug use, psychiatric symptoms, antisocial behavior, and disinhibited personality traits among a nationally representative survey of U.S. adults. We were especially interested to know what characteristics differed between people with CHS symptoms and those who use cannabis daily but without CHS symptoms, though we also examined difference with people with less frequent cannabis use to provide a reference similar to healthy controls. These findings will help shape an understanding of the psychological and behavioral profile of people experiencing CHS symptoms and their unique needs to improve diagnostic and intervention approaches.

## 2. METHODS

### 2.1. Study design

Data were collected as part of the National Firearms, Alcohol, Cannabis, and Suicide (NFACS) survey (*N* = 7,034) nationally representative survey of U.S. adults, the details of which have been described elsewhere. (Ilgen, 2026) The survey was developed by the study team and fielded by a market research company, Verasight, from May 27 – September 2, 2025. Participants were recruited via random address-based sampling (ABS) or MMS and SMS text message methods. For the ABS sample, potential participants were identified through the U.S. Postal Service database and mailed an initial invitation letter as well as a reminder postcard.

Where corresponding cell phone numbers and email addresses were available, the research company also sent follow-up reminders with a direct URL to the web-based survey utilizing those alternative contact methods. For the MMS and/or SMS text message recruitment, potential participants were identified from a database consisting of voter file and supplemental commercial records and received a survey link via text message. Response rates were 3.83% (*n* = 1,914) and 0.39% (*n* = 8,652) for the ABS and MAMS/SMS text messaging, respectively. From these responses, 2,829 (26.7%) were removed due to incomplete data. An additional 703 responses (6.7%) were excluded during the quality assurance process, which included confirming all responses corresponded with a U.S. IP address, no duplicate responses were provided, no non-human responses were included and removing participants who completed the survey in less than 30% of the median completion time. This yielded a sample of 1,509 ABS-recruited and 5,525 MMS/SMS-recruited participants. Though the response rate may seem low, these rates are consistent with participation rates when establishing new cohorts rather existing panels,(Dutwin, 2024) and the low response rates do not violate the assumptions of probability-based sampling, which are that all those invited have an equal probability of participating. Raking was used to calculate poststratification weights to match the sample characteristics to national benchmarks from the July 2025 Current Population Survey for age, sex, race/ethnicity, income, education, political party identification, region, and metropolitan status, so that inferences using the sample data better generalize to the target population of US adults.

The protocol was reviewed by the University of Michigan IRB and received an exempt determination. Prior to accessing the survey, participants reviewed information on the survey content, length of the survey and compensation, and were informed that participation was voluntary and anonymous. All participants provided active consent prior to receiving survey questions and all participants completed the survey online.

### 2.2. Measures

#### 2.2.1. CHS symptoms

We asked a series of questions to capture a group of participants whose response profile was consistent with CHS. All respondents who endorsed at least 20 lifetime uses of cannabis were asked if they had a period of daily or near daily cannabis use in the past 5 years. Those who endorsed daily use in the past 5 years were asked if they had experienced periods of severe nausea, vomiting, or abdominal pain. Using these responses, we made four groups for comparisons: the CHS symptom group included respondents who endorsed a period of daily cannabis use in the past 5 years and periods of nausea, vomiting, or abdominal pain (*n* = 191); all other respondents who endorsed a period of daily cannabis use in the past 5 years but not periods of severe nausea, vomiting, or abdominal pain (*n* = 882); all other respondents who reported cannabis use in the past 12 months (*n* = 1288); all other respondents who we refer to as the no cannabis use in the past 12 months group (*n* = 4673).

#### 2.2.2. Drug and alcohol use

Lifetime and past 12 month alcohol, nicotine, and drug use was assessed using the Alcohol, Smoking, and Substance Involvement Screening Test (ASSIST). (Group, 2002) Negative consequences of alcohol use were assessed using items adapted from the Alcohol Use Disorders Identification Test (AUDIT) (Saunders et al., 1993) with some additions including items that assessed lifetime problem use as well as past 12 month prevalence. We also asked if the participants had experienced a drug overdose or alcohol poisoning in their lifetime and in the past 12 months.

#### 2.2.3. Mental health problems and social support

We used the PHQ-2 (α = .88) (Kroenke et al., 2003) and two items from the GAD-2 (α = .87) (Kroenke et al., 2007) to assess depression and generalized anxiety over the past 2 weeks, respectively. We also included five items from the National Institute of Health (NIH) Toolbox Adult Social Relationship Scales (Cyranowski et al., 2013) that assessed perceived social support (e.g., people available for emotional and physical support; α = .80).

#### 2.2.4. Sleep problems, pain, and overall health

Participants completed four items on sleep problems (e.g., trouble falling asleep, staying asleep, waking up; α = .87) over the past 30 days. (Jenkins et al., 1988) Participants also completed the Pain, Enjoyment of Life, General Activity (PEG) scale, containing three items that rated their level of pain over the past week and how much this pain interfered with their enjoyment of life and activity level (α = .84). (Krebs et al., 2009) Participants also rated their overall health (poor, fair, good, very good, excellent). (Ware et al., 1996)

#### 2.2.5. Antisocial behavior

Participants completed seven items that asked how many times (*never, once or twice, several times, many times*) they had engaged in various types of antisocial behavior including theft, destroying property, lying and conning, and aggression and violence, as well as being arrested and suspended or expelled from school (α = .79).

#### 2.2.6. Intimate partner violence

Participants were also asked if they ever had a romantic relationship with the same partner that lasted longer than 1 month. If yes (*n* = 6372), participants completed 5 items adapted from the Controlling and Abusive Tactics-2 (Hamel et al., 2015) and Conflict Tactics Scale-2 (STRAUS et al., 1996) that asked how frequently (*never, rare, occasional, common, frequent*) they attempt to control who their current or most recent romantic partner spends time with; search their partner’s purse, wallet, or cell phone; threaten to hurt their partner; push, slap, hit, kick, punch or choke their partner; use a knife, gun, or other weapon on their partner (α = .63). Participants were also asked if they had ever been the victim of the same behaviors at the hand of their intimate partner (α = .84).

#### 2.2.7. Personality

We included markers of several personality traits including antagonism vs agreeableness (3 items, α = .72) and conscientiousness (4 items, α = .62) (Soto & John, 2017); negative urgency (4 items, α = .83) (Cyders et al., 2014); persecutory ideas (3 items, α = .72); desire for power (3 items, α = .71) and feeling powerless (3 items, α = .72) (Murphy et al., 2022); and subjective well-being (4 items, α = .84). (Salsman et al., 2020)

### 2.3. Analyses

Logistic and linear regression models were fit to compare the CHS symptom and other cannabis use groups on categorical and quantitative measures, respectively, of substance use, mental health, and personality variables. Our first goal was to determine if the CHS symptom group differed from the daily cannabis use group as this would suggest a high-risk sub-group with additional psychological, behavioral, and health problems. We also compared the CHS symptom and daily use groups to the less-than-daily cannabis use groups to provide a broader context for their level of psychosocial functioning relative to low drug using groups more comparable to healthy controls. Analyses were conducted in Stata 19 using the Survey data analysis module with poststratification weights to adjust parameter estimates and standard errors.

## 3. RESULTS

### 3.1. Drug use

Table 1 presents the lifetime prevalence rates and group differences for drug overdoses and use of various classes of drugs. The CHS symptom group had higher lifetime rates of drug overdose than all the other groups, and those with daily use had higher rates of drug overdose than the no cannabis use in the past 12 months group. The CHS symptom and daily use groups also had higher lifetime rates of alcohol poisoning than the no cannabis use in the past 12 months group but did not differ from each other.

**Table 1.** Lifetime prevalence rates of drug overdoses and drug use among CHS and cannabis use groups.

| Drug use and problem use | CHS symptoms<br>(n = 191)<sup>a</sup> | Daily cannabis use in<br>the past 5 years<br>(n = 882)<sup>b</sup> | Cannabis use<br>past 12 months<br>(n = 1288)<sup>c</sup> | No cannabis use<br>past 12 months<br>(n = 4673)<sup>d</sup> |
| --- | --- | --- | --- | --- |
| Drug overdose lifetime |  |  |  |  |
| % (n) | 21.0 (40)<sup>bcd</sup> | 7.4 (66)<sup>ad</sup> | 5.1 (65)<sup>ad</sup> | 1.9 (90)<sup>abc</sup> |
| OR | **3.32\*\*\* (1.71, 6.44)** | ref | 0.66 (0.39, 1.12) | **0.24\*\*\* (0.15, 0.39)** |
| Alcohol poisoning lifetime |  |  |  |  |
| % (n) | 18.7 (36)<sup>d</sup> | 12.2 (108)<sup>d</sup> | 7.3 (94)<sup>d</sup> | 3.0 (142)<sup>abc</sup> |
| OR | 1.65 (0.89, 3.07) | ref | **0.57\*\* (0.38, 0.85)** | **0.23\*\*\* (0.16, 0.32)** |
| Binge drinking episode lifetime |  |  |  |  |
| % (n) | 72.7 (139)<sup>cd</sup> | 69.5 (612)<sup>cd</sup> | 57.1 (735)<sup>abd</sup> | 36.1 (1689)<sup>abc</sup> |
| OR | 1.17 (0.68, 2.01) | ref | **0.58\*\*\* (0.45, 0.75)** | **0.25\*\*\* (0.20, 0.31)** |
| Hallucinogens or psychedelics lifetime |  |  |  |  |
| % (n) | 71.9 (137)<sup>bcd</sup> | 53.5 (471)<sup>acd</sup> | 26.3 (339)<sup>abd</sup> | 8.6 (402)<sup>abc</sup> |
| OR | **2.23\*\* (1.35, 3.68)** | ref | **0.31\*\*\* (0.24, 0.40)** | **0.08 \*\*\* (0.07, 0.10)** |
| Cocaine lifetime |  |  |  |  |
| % (n) | 49.7 (95)<sup>cd</sup> | 44.3 (391)<sup>cd</sup> | 23.5 (302)<sup>abd</sup> | 9.0 (420)<sup>abc</sup> |
| OR | 1.24 (0.78, 1.98) | ref | **0.39\*\*\* (0.30, 0.49)** | **0.12\*\*\* (0.10, 0.16)** |
| Sedatives or sleeping pills lifetime |  |  |  |  |
| % (n) | 49.3 (94)<sup>cd</sup> | 36.5 (322)<sup>cd</sup> | 25.5 (328)<sup>abd</sup> | 12.3 (576)<sup>abc</sup> |
| OR | **1.69\* (1.06, 2.70)** | ref | **0.59\*\*\* (0.46, 0.76)** | **0.24\*\*\* (0.20, 0.31)** |
| Prescription opioids lifetime |  |  |  |  |
| % (n) | 47.0 (90)<sup>cd</sup> | 39.0 (344)<sup>cd</sup> | 27.7 (357)<sup>abd</sup> | 19.1 (892)<sup>abc</sup> |
| OR | 1.39 (0.87, 2.21) | ref | **0.60\*\*\* (0.47, 0.77)** | **0.37\*\*\* (0.30, 0.46)** |
| Amphetamine-type stimulants lifetime |  |  |  |  |
| % (n) | 42.6 (81)<sup>cd</sup> | 42.8 (377)<sup>cd</sup> | 24.5 (315)<sup>abd</sup> | 9.9 (461)<sup>abc</sup> |
| OR | 1.00 (0.63, 1.57) | ref | **0.43\*\*\* (0.34, 0.56)** | **0.15\*\*\* (0.12, 0.18)** |
| Kratom lifetime |  |  |  |  |
| % (*n*) | 21.5 (41)<sup>cd</sup> | 10.8 (95)<sup>cd</sup> | 4.0 (52)<sup>abd</sup> | 1.4 (67)<sup>abc</sup> |
| OR | **2.25\* (1.19, 4.28)** | ref | **0.35\*\*\* (0.20, 0.55)** | **0.12\*\*\* (0.08, 0.19)** |
| Heroin or fentanyl lifetime |  |  |  |  |
| % (*n*) | 20.9 (40)<sup>bcd</sup> | 8.3 (73)<sup>acd</sup> | 4.2 (54)<sup>abd</sup> | 1.8 (84)<sup>abc</sup> |
| OR | **2.93\*\* (1.49, 5.78)** | ref | **0.48\*\* (0.29, 0.82)** | **0.20\*\*\* (0.13, 0.33)** |
| Inhalants lifetime |  |  |  |  |
| % (*n*) | 16.6 (32)<sup>cd</sup> | 13.8 (121)<sup>cd</sup> | 6.7 (87)<sup>abd</sup> | 2.7 (128)<sup>abc</sup> |
| OR | 1.24 (0.68, 2.28) | ref | **0.45\*\*\* (0.30, 0.68)** | **0.18\*\*\* (0.12, 0.26)** |

Note. \* $p < .05$ ; \*\* $p < .01$ ; \*\*\* $p < .001$ . OR = odds ratio; CHS = Cannabinoid hyperemesis syndrome. The daily cannabis use refers to respondents who endorsed daily or near daily cannabis use in the past 5 years who did not report CHS symptoms. Subscripts indicate a significant difference between groups. Pairwise comparisons were judged to be statistically significant if the 95% confidence intervals for Bonferroni adjusted post hoc tests did not include zero.

The CHS symptom group had the highest lifetime rates of use for nearly every drug class, though not all the group comparisons were statistically significant. Compared to the daily cannabis use group, the CHS symptom group had significantly higher lifetime rates of use of heroin or fentanyl, hallucinogens or psychedelics, sedatives or sleeping pills, and kratom, though the latter two pairwise comparisons were not significant after the Bonferroni adjustment for post hoc tests. Compared to the two lower-level cannabis use groups, the CHS symptom and daily use groups had higher lifetime rates of use for each drug class and binge drinking.

The CHS symptom and daily use groups reported trying a greater variety of drugs and more lifetime alcohol use problems than the less-than-daily cannabis use groups but did not differ from each other (see Table 3). Those with cannabis use reported higher rates of use for each drug class, using a greater variety of drugs, and more lifetime alcohol use problems than the no cannabis use.

Table 2 presents the prevalence rates and group differences for use of various types of drugs in the past 12 months. The CHS symptom group had higher rates of drug overdoses and alcohol poisoning in the past 12 months than all the other groups, but the prevalence of each was so low these differences may be unreliable. Compared to the daily and cannabis use in the past 12 months groups, the CHS symptom group had higher rates of use of heroin or fentanyl, prescription opioids, and sedatives or sleeping pills in the past 12 months, though the pairwise comparisons were not significant after adjusting for multiple comparisons. Compared to the cannabis use in the past 12 months group, the CHS symptom and daily use groups had higher rates of daily nicotine use and use of hallucinogens or psychedelics in the past 12 months.

**Table 2.** Past 12 month prevalence rates of drug overdoses and drug and alcohol use among CHS and cannabis use groups.

| Drug use and problem use | CHS symptoms<br>(n = 191)<sup>a</sup> | Daily cannabis use in<br>the past 5 years<br>(n = 882)<sup>b</sup> | Cannabis use<br>past 12 months<br>(n = 1288)<sup>c</sup> | No cannabis use<br>past 12 months<br>(n = 4673)<sup>d</sup> |
| --- | --- | --- | --- | --- |
| Drug overdose past 12 months |  |  |  |  |
| % (n) | 4.8 (9) | 0.9 (8) | 1.1 (14) | 0.1 (6) |
| OR | 5.49 (0.96, 31.3) | ref | 1.18 (0.23, 5.9) | **0.14\* (0.02, 0.80)** |
| Alcohol poisoning past 12 months |  |  |  |  |
| % (n) | 6.1 (12) | 0.1 (1) | 0.4 (5) | 0.1 (5) |
| OR | **85.8\*\*\* (9.37, 785.3)** | ref | 5.69 (0.67, 48.4) | 1.46 (0.18, 11.7) |
| Daily nicotine use past 12 months |  |  |  |  |
| % (n) | 44.5 (72)<sup>cd</sup> | 36.0 (268)<sup>cd</sup> | 19.7 (248)<sup>abd</sup> | 11.3 (445)<sup>abc</sup> |
| OR | 1.43 (0.89, 2.28) | ref | **0.44\*\*\* (0.33, 0.57)** | **0.23\*\*\* (0.18, 0.29)** |
| Binge drinking episode past 30 days |  |  |  |  |
| % (n) | 30.0 (41)<sup>d</sup> | 30.1 (223)<sup>d</sup> | 26.1 (356)<sup>d</sup> | 10.9 (498)<sup>abc</sup> |
| OR | 0.99 (0.59, 1.67) | ref | 0.82 (0.63, 1.07) | **0.28\*\*\* (0.22, 0.36)** |
| Heavy alcohol use past 12 months |  |  |  |  |
| % (n) | 17.9 (25)<sup>d</sup> | 14.1 (89)<sup>d</sup> | 10.6 (137)<sup>d</sup> | 3.9 (167)<sup>abc</sup> |
| OR | 1.33 (0.68, 2.57) | ref | 0.72 (0.50, 1.05) | **0.25\*\*\* (0.18, 0.35)** |
| Daily alcohol use past 12 months |  |  |  |  |
| % (n) | 17.0 (23)<sup>d</sup> | 10.8 (109)<sup>d</sup> | 11.6 (190)<sup>d</sup> | 4.8 (279)<sup>abc</sup> |
| OR | 1.69 (0.91, 3.14) | ref | 1.08 (0.78, 1.49) | **0.42\*\*\* (0.31, 0.56)** |
| Hallucinogens or Psychedelics past 12 months |  |  |  |  |
| % (n) | 33.0 (63)<sup>cd</sup> | 25.6 (226)<sup>cd</sup> | 9.8 (126)<sup>abd</sup> | 0.9 (43)<sup>abc</sup> |
| OR | 1.43 (0.85, 2.41) | ref | **0.32\*\*\* (0.23, 0.43)** | **0.03\*\*\* (0.02, 0.04)** |
| Sedatives or sleeping pills past 12 months |  |  |  |  |
| % (n) | 27.9 (53)<sup>d</sup> | 17.2 (152)<sup>d</sup> | 15.4 (198)<sup>d</sup> | 6.6 (310)<sup>abc</sup> |
| OR | **1.86\* (1.08, 3.22)** | ref | 0.87 (0.64, 1.19) | **0.34\*\*\* (0.26, 0.45)** |
| Prescription opioids past 12 months |  |  |  |  |
| % (n) | 26.1 (50)<sup>d</sup> | 14.8 (131)<sup>d</sup> | 15.1 (194)<sup>d</sup> | 6.9 (325)<sup>abc</sup> |
| OR | **2.03\* (1.17, 3.52)** | ref | 1.02 (0.74, 1.40) | **0.43\*\*\* (0.32, 0.57)** |
| Amphetamine-type stimulants past 12 months |  |  |  |  |
| % (n) | 20.0 (38)<sup>d</sup> | 17.7 (156)<sup>cd</sup> | 11.1 (143)<sup>bd</sup> | 2.4 (114)<sup>abc</sup> |
| OR | 1.17 (0.66, 2.07) | ref | **0.58\*\* (0.41, 0.81)** | **0.12\*\*\* (0.08, 0.17)** |
| Cocaine past 12 months |  |  |  |  |
| % (n) | 18.8 (36)<sup>d</sup> | 14.9 (131)<sup>cd</sup> | 7.7 (99)<sup>bd</sup> | 0.7 (31)<sup>abc</sup> |
| OR | 1.32 (0.69, 2.51) | ref | **0.48\*\*\* (0.33, 0.69)** | **0.04\*\*\* (0.02, 0.07)** |
| Kratom past 12 months |  |  |  |  |
| % (n) | 9.3 (18) | 5.2 (46)<sup>d</sup> | 2.3 (29)<sup>d</sup> | 0.5 (25)<sup>bc</sup> |
| OR | 1.88 (0.71, 5.01) | ref | **0.42\*\* (0.22, 0.81)** | **0.10\*\*\* (0.5, 0.21)** |
| Heroin or Fentanyl past 12 months |  |  |  |  |
| % (n) | 8.4 (16) | 0.9 (8) | 1.6 (20)<sup>d</sup> | 0.3 (12)<sup>c</sup> |
| OR | **10.5\*\*\* (2.94, 37.6)** | ref | 1.82 (0.64, 5.17) | **0.29\* (0.09, 0.995)** |
| Inhalants past 12 months |  |  |  |  |
| % (n) | 5.1 (10) | 3.9 (35)<sup>d</sup> | 2.1 (27)<sup>d</sup> | 0.3 (15)<sup>abc</sup> |
| OR | 1.31 (0.36, 4.73) | ref | 0.53 (0.25, 1.12) | **0.08\*\*\* (0.03, 0.24)** |

Note. \* $p < .05$ ; \*\* $p < .01$ ; \*\*\* $p < .001$ . OR = odds ratio; CHS = Cannabinoid hyperemesis syndrome. The daily cannabis use refers to respondents who endorsed daily or near daily cannabis use in the past 5 years who did not report CHS symptoms. Subscripts indicate a significant difference between groups. Pairwise comparisons were judged to be statistically significant if the 95% confidence intervals for Bonferroni adjusted post hoc tests did not include zero.

**Table 3.**
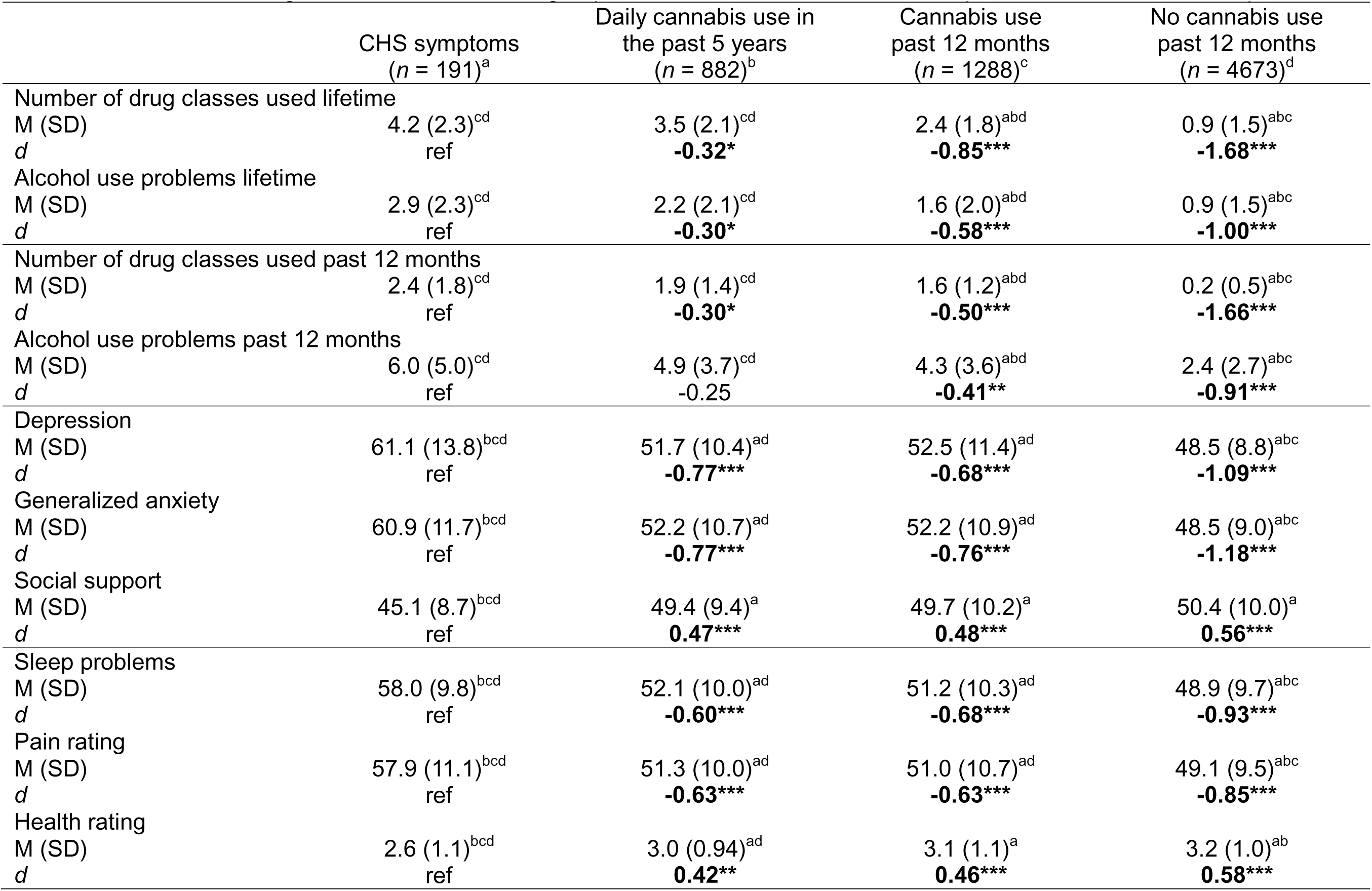
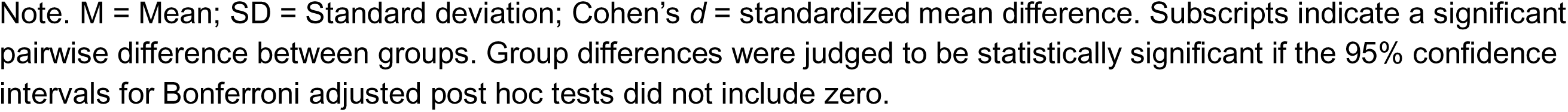
Differences among CHS and cannabis use groups on substance use, mental health problems, and other health problems.

|  | CHS symptoms<br>(*n* = 191)<sup>a</sup> | Daily cannabis use in<br>the past 5 years<br>(*n* = 882)<sup>b</sup> | Cannabis use<br>past 12 months<br>(*n* = 1288)<sup>c</sup> | No cannabis use<br>past 12 months<br>(*n* = 4673)<sup>d</sup> |
| --- | --- | --- | --- | --- |
| Number of drug classes used lifetime |  |  |  |  |
| M (SD) | 4.2 (2.3)<sup>cd</sup> | 3.5 (2.1)<sup>cd</sup> | 2.4 (1.8)<sup>abd</sup> | 0.9 (1.5)<sup>abc</sup> |
| *d* | ref | **-0.32\*** | **-0.85\*\*\*** | **-1.68\*\*\*** |
| Alcohol use problems lifetime |  |  |  |  |
| M (SD) | 2.9 (2.3)<sup>cd</sup> | 2.2 (2.1)<sup>cd</sup> | 1.6 (2.0)<sup>abd</sup> | 0.9 (1.5)<sup>abc</sup> |
| *d* | ref | **-0.30\*** | **-0.58\*\*\*** | **-1.00\*\*\*** |
| Number of drug classes used past 12 months |  |  |  |  |
| M (SD) | 2.4 (1.8)<sup>cd</sup> | 1.9 (1.4)<sup>cd</sup> | 1.6 (1.2)<sup>abd</sup> | 0.2 (0.5)<sup>abc</sup> |
| *d* | ref | **-0.30\*** | **-0.50\*\*\*** | **-1.66\*\*\*** |
| Alcohol use problems past 12 months |  |  |  |  |
| M (SD) | 6.0 (5.0)<sup>cd</sup> | 4.9 (3.7)<sup>cd</sup> | 4.3 (3.6)<sup>abd</sup> | 2.4 (2.7)<sup>abc</sup> |
| *d* | ref | -0.25 | **-0.41\*\*** | **-0.91\*\*\*** |
| Depression |  |  |  |  |
| M (SD) | 61.1 (13.8)<sup>bcd</sup> | 51.7 (10.4)<sup>ad</sup> | 52.5 (11.4)<sup>ad</sup> | 48.5 (8.8)<sup>abc</sup> |
| *d* | ref | **-0.77\*\*\*** | **-0.68\*\*\*** | **-1.09\*\*\*** |
| Generalized anxiety |  |  |  |  |
| M (SD) | 60.9 (11.7)<sup>bcd</sup> | 52.2 (10.7)<sup>ad</sup> | 52.2 (10.9)<sup>ad</sup> | 48.5 (9.0)<sup>abc</sup> |
| *d* | ref | **-0.77\*\*\*** | **-0.76\*\*\*** | **-1.18\*\*\*** |
| Social support |  |  |  |  |
| M (SD) | 45.1 (8.7)<sup>bcd</sup> | 49.4 (9.4)<sup>a</sup> | 49.7 (10.2)<sup>a</sup> | 50.4 (10.0)<sup>a</sup> |
| *d* | ref | **0.47\*\*\*** | **0.48\*\*\*** | **0.56\*\*\*** |
| Sleep problems |  |  |  |  |
| M (SD) | 58.0 (9.8)<sup>bcd</sup> | 52.1 (10.0)<sup>ad</sup> | 51.2 (10.3)<sup>ad</sup> | 48.9 (9.7)<sup>abc</sup> |
| *d* | ref | **-0.60\*\*\*** | **-0.68\*\*\*** | **-0.93\*\*\*** |
| Pain rating |  |  |  |  |
| M (SD) | 57.9 (11.1)<sup>bcd</sup> | 51.3 (10.0)<sup>ad</sup> | 51.0 (10.7)<sup>ad</sup> | 49.1 (9.5)<sup>abc</sup> |
| *d* | ref | **-0.63\*\*\*** | **-0.63\*\*\*** | **-0.85\*\*\*** |
| Health rating |  |  |  |  |
| M (SD) | 2.6 (1.1)<sup>bcd</sup> | 3.0 (0.94)<sup>ad</sup> | 3.1 (1.1)<sup>a</sup> | 3.2 (1.0)<sup>ab</sup> |
| *d* | ref | **0.42\*\*** | **0.46\*\*\*** | **0.58\*\*\*** |

Note. M = Mean; SD = Standard deviation; Cohen's $d$ = standardized mean difference. Subscripts indicate a significant pairwise difference between groups. Group differences were judged to be statistically significant if the 95% confidence intervals for Bonferroni adjusted post hoc tests did not include zero.

Compared to the no cannabis use in the past 12 months group, the CHS symptom, daily use, and cannabis use in the past 12 months groups had higher rates of daily nicotine and alcohol use, heavy alcohol use, and each drug class in the past 12 months as well as binge drinking in the past 30 days. The CHS symptom and daily use groups reported using a greater variety of drugs and more alcohol use problems in the past 12 months than the less-than-daily and no cannabis use in past 12 months groups but did not differ from each other (see Table 3). The cannabis use in the past 12 months group also reported using a greater variety of drugs and more alcohol use problems in the past 12 months than the no cannabis use group.

### 3.2. Mental health problems, sleep problems, pain, and overall health

For the remaining group comparisons, differences with the CHS symptom group tended to be of large effect size for the no cannabis use in the past 12 months group and medium-to-large for the daily and less-than-daily cannabis use groups. The daily and less-than-daily cannabis use groups tended not to exhibit significant differences from each other and to have small-to-medium sized differences with the no cannabis use in the past 12 months group that were in the same direction as the CHS symptom group.

Table 3 presents the group differences for mental health problems and other health-related variables. The CHS symptom group reported higher levels of depression (mean *d* = 0.84) and generalized anxiety in the past two weeks (mean *d* = 0.90) and lower levels of social support (mean *d* = −0.50) than all the other groups. The CHS symptom group also reported more sleep problems (mean *d* = 0.73), higher pain ratings (mean *d* = 0.70), and lower overall health ratings (mean *d* = −0.49) than all the other groups.

### 3.3. Antisocial behavior and intimate partner violence

The CHS symptom group reported higher rates of antisocial behavior (mean *d* = 0.92) and being both the perpetrator (mean *d* = 0.71) and victim (mean *d* = 0.73) of intimate partner violence (see Table 4) than all the other groups.

**Table 4.**
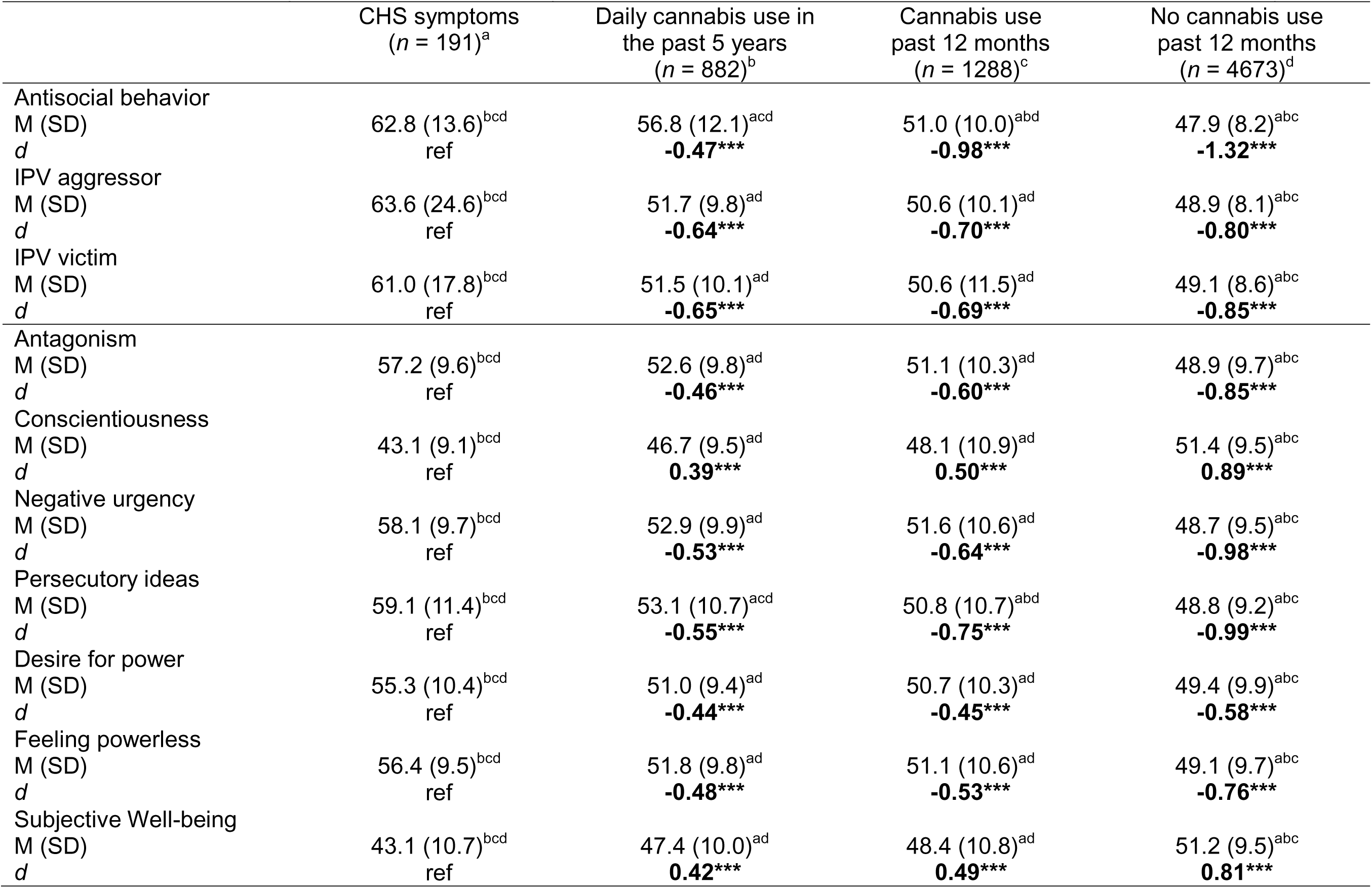
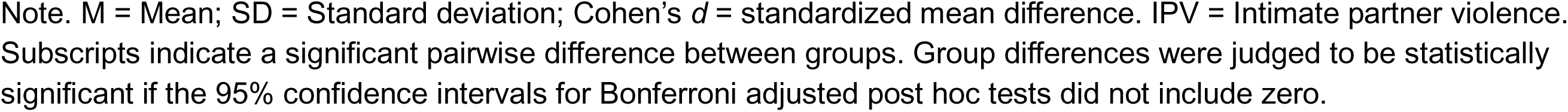
Differences among CHS and cannabis use groups on antisocial behavior, intimate partner violence, and personality.

|  | CHS symptoms<br>(*n* = 191)<sup>a</sup> | Daily cannabis use in<br>the past 5 years<br>(*n* = 882)<sup>b</sup> | Cannabis use<br>past 12 months<br>(*n* = 1288)<sup>c</sup> | No cannabis use<br>past 12 months<br>(*n* = 4673)<sup>d</sup> |
| --- | --- | --- | --- | --- |
| Antisocial behavior |  |  |  |  |
| M (SD) | 62.8 (13.6)<sup>bcd</sup> | 56.8 (12.1)<sup>acd</sup> | 51.0 (10.0)<sup>abd</sup> | 47.9 (8.2)<sup>abc</sup> |
| *d* | ref | **-0.47\*\*\*** | **-0.98\*\*\*** | **-1.32\*\*\*** |
| IPV aggressor |  |  |  |  |
| M (SD) | 63.6 (24.6)<sup>bcd</sup> | 51.7 (9.8)<sup>ad</sup> | 50.6 (10.1)<sup>ad</sup> | 48.9 (8.1)<sup>abc</sup> |
| *d* | ref | **-0.64\*\*\*** | **-0.70\*\*\*** | **-0.80\*\*\*** |
| IPV victim |  |  |  |  |
| M (SD) | 61.0 (17.8)<sup>bcd</sup> | 51.5 (10.1)<sup>ad</sup> | 50.6 (11.5)<sup>ad</sup> | 49.1 (8.6)<sup>abc</sup> |
| *d* | ref | **-0.65\*\*\*** | **-0.69\*\*\*** | **-0.85\*\*\*** |
| Antagonism |  |  |  |  |
| M (SD) | 57.2 (9.6)<sup>bcd</sup> | 52.6 (9.8)<sup>ad</sup> | 51.1 (10.3)<sup>ad</sup> | 48.9 (9.7)<sup>abc</sup> |
| *d* | ref | **-0.46\*\*\*** | **-0.60\*\*\*** | **-0.85\*\*\*** |
| Conscientiousness |  |  |  |  |
| M (SD) | 43.1 (9.1)<sup>bcd</sup> | 46.7 (9.5)<sup>ad</sup> | 48.1 (10.9)<sup>ad</sup> | 51.4 (9.5)<sup>abc</sup> |
| *d* | ref | **0.39\*\*\*** | **0.50\*\*\*** | **0.89\*\*\*** |
| Negative urgency |  |  |  |  |
| M (SD) | 58.1 (9.7)<sup>bcd</sup> | 52.9 (9.9)<sup>ad</sup> | 51.6 (10.6)<sup>ad</sup> | 48.7 (9.5)<sup>abc</sup> |
| *d* | ref | **-0.53\*\*\*** | **-0.64\*\*\*** | **-0.98\*\*\*** |
| Persecutory ideas |  |  |  |  |
| M (SD) | 59.1 (11.4)<sup>bcd</sup> | 53.1 (10.7)<sup>acd</sup> | 50.8 (10.7)<sup>abd</sup> | 48.8 (9.2)<sup>abc</sup> |
| *d* | ref | **-0.55\*\*\*** | **-0.75\*\*\*** | **-0.99\*\*\*** |
| Desire for power |  |  |  |  |
| M (SD) | 55.3 (10.4)<sup>bcd</sup> | 51.0 (9.4)<sup>ad</sup> | 50.7 (10.3)<sup>ad</sup> | 49.4 (9.9)<sup>abc</sup> |
| *d* | ref | **-0.44\*\*\*** | **-0.45\*\*\*** | **-0.58\*\*\*** |
| Feeling powerless |  |  |  |  |
| M (SD) | 56.4 (9.5)<sup>bcd</sup> | 51.8 (9.8)<sup>ad</sup> | 51.1 (10.6)<sup>ad</sup> | 49.1 (9.7)<sup>abc</sup> |
| *d* | ref | **-0.48\*\*\*** | **-0.53\*\*\*** | **-0.76\*\*\*** |
| Subjective Well-being |  |  |  |  |
| M (SD) | 43.1 (10.7)<sup>bcd</sup> | 47.4 (10.0)<sup>ad</sup> | 48.4 (10.8)<sup>ad</sup> | 51.2 (9.5)<sup>abc</sup> |
| *d* | ref | **0.42\*\*\*** | **0.49\*\*\*** | **0.81\*\*\*** |

Note. M = Mean; SD = Standard deviation; Cohen's $d$ = standardized mean difference. IPV = Intimate partner violence. Subscripts indicate a significant pairwise difference between groups. Group differences were judged to be statistically significant if the 95% confidence intervals for Bonferroni adjusted post hoc tests did not include zero.

### 3.4. Personality

The CHS symptom group exhibited significant differences from all the other groups on each personality trait, and all the effects were in the direction of worse psychosocial functioning. The CHS symptom group endorsed significantly higher antagonism (mean *d* = 0.64), negative urgency (mean *d* = 0.72), persecutory ideas (mean *d* = 0.76), desire for power (mean *d* = 0.48), and feeling powerless (mean *d* = 0.59), and lower conscientiousness (mean *d* = −0.59) and subjective well-being (mean *d* = −0.57) than all the other groups.

### 3.5. Supplemental analyses

As described in the Supplemental materials, we also conducted the analyses three additional ways using narrower criteria for the CHS symptom group: (1) vomiting episodes must occur at least 1 x month (*n* = 141), (2) only include respondents who report symptom relief from hot baths/showers or prolonged cessation of cannabis use (*n* = 105), (3) only include those who meet criteria for 1 and 2 (*n* = 80). Across these methods, we found that each CHS symptom group exhibited a similar pattern of differences with the other cannabis use groups as did the CHS symptom group in the primary analyses, including similar effect sizes for the comparisons (however, some pairwise comparisons were no longer statistically significant due to the smaller sample sizes of the alternative CHS symptom groups). Therefore, we identified a similar group in terms of their psychological and behavioral characteristics regardless of how wide or narrow the criteria for inclusion in the CHS symptom group.

## 4. DISCUSSON

CHS is an emerging disorder as the accessibility and rates of heavy cannabis use increase in the United States. We previously showed that almost 18% of those who report daily cannabis use also reported symptoms consistent with CHS (i.e., episodes of severe nausea, vomiting, and abdominal pain), and these people exhibited more symptoms of cannabis use disorder than those who use cannabis daily but do not have CHS symptoms.(Ilgen, 2026) We expanded these findings by showing that the CHS symptom group reported greater substance use, anxiety and depressive symptoms, antisocial behavior, and personality characteristics associated with greater social problems, even when compared to the group of individuals who used cannabis daily but did not report CHS symptoms.

CHS symptoms and daily cannabis use was associated with a greater variety of drug use relative to those with less-than-daily levels of cannabis use. There was some evidence that those with CHS symptoms had higher rates of opioid and sedative use compared to others who used cannabis daily, suggesting CHS symptoms may have an especially strong association with “downer” drugs that have sedative effects, but not “upper” drugs that have stimulant effects. A question for future research is whether this pattern of drug use is related to attempts to relieve CHS symptoms, comorbid or related conditions (e.g., more sleep problems and chronic pain), or is attributable to common pre-morbid risk factors. CHS symptoms were also associated with a significantly higher rate of lifetime drug overdoses which may be related to their high rates of opioid and sedative use and alcohol use problems and the potential for drug and alcohol co-use. Those with CHS symptoms reported higher levels of alcohol use and problem than those had not used cannabis in the past 12 months but had similar levels of alcohol use compared to others who use cannabis.

Those with CHS symptoms were elevated on several mental health (depression, anxiety) and health-related problems (poor sleep, pain, worse overall health) relative to all the other groups and reported the lowest levels of social support. People with CHS symptoms also reported the highest levels of antisocial behavior and intimate partner violence and a personality profile associated with more externalizing problems (e.g. antagonism, negative urgency, low conscientiousness). The CHS symptom group also reported the lowest subjective well-being and the most feelings of powerlessness, which dovetailed with their high levels of mental health and other health-related problems. While the CHS symptom group was different from the daily cannabis use group on all these attributes, they exhibited extreme differences when compared to those who did not use cannabis in the past 12 months which may provide a better referent to illustrate the breadth and depth of their psychosocial challenges.

As the prevalence of CHS grows, it is becoming increasingly important to develop cohesive and effective strategies for managing CHS. Because CHS is often treated in acute care settings, much of the emphasis has been on diagnosis and the short-term management of CHS symptoms (e.g., the American Gastroenterological Association (AGA) Clinical Practice Update) (Rubio-Tapia et al., 2024). Beyond the acute management of nausea and vomiting, people with CHS symptoms also exhibit significant substance- and mental health-related problems. Parallel approaches will be needed to develop strategies needed to support these individuals in making longer-lasting changes to their substance use. Given the general link between psychopathology and poorer substance use disorder treatment outcomes, those with CHS symptoms may struggle with successfully changing their cannabis use and may require a coordinated approach to deal with co-occurring substance use and mental health symptoms (Iqbal et al., 2019).

The most significant limitation of the design was the inability to directly attribute the nausea and vomiting in the CHS symptom group to cannabis use. Our inclusion criteria for the CHS symptom group prioritized sensitivity over specificity, and it is possible that some in the CHS symptom group experienced gastrointestinal symptoms for reasons unrelated to cannabis use. This wide-net strategy should reduce differences between the CHS symptom group and other groups based on patterns of cannabis use. Despite using these non-specific inclusion criteria, we still detected differences between those reporting CHS symptoms and others cannabis groups on multiple measures of substance use and poor psychosocial functioning. To further address this concern, we conducted multiple supplemental analyses using increasingly narrow criteria for inclusion into the CHS symptom group (e.g., at least 1 vomiting episode a month, symptom relief from hot baths/showers and prolonged cessation of cannabis use, or both) and obtained very similar results. Thus, whether focusing only on the essential features of CHS or narrowing the CHS symptom group to be more severe and prototypical, we identified groups that were very similar in their psychological and behavioral profile. Future work will need to focus on improved assessment of CHS symptoms and differential diagnosis, but our results consistently find that those with CHS symptoms experience significant substance use and mental health problems.

Other limitations include that while the overall sample was large, the low prevalence of many drug-related outcomes–especially for use in the past 12 months–made it difficult to detect significant group differences. Similarly, the survey provided broad assessments of numerous domains but often lacked detail about the extent of prior substance use (e.g., age of onset, years of use) and other mental health symptoms. Also, establishing the links among CHS symptoms, psychiatric symptoms, and substance use does not reveal the causal nature of the relationships. For example, psychiatric symptoms may be a risk factor, comorbid condition, or consequence of a recurrent illness. Research designs that allow for temporal ordering of the relationships will be needed to make these distinctions. The survey design also required participants to have online access and proficiency in English which may have limited generalizability. Although survey weights were utilized to better approximate the adult US population, they may have failed to adjust for other unmeasured sources of bias.

Our results indicate that adults with CHS symptoms exhibit not only elevated levels of cannabis use problems but also more mental problems, heavy drug use, antisocial behavior, dysfunctional personality traits and other health-related problems, relative to those with daily cannabis use and differences that are quite sizable when compared to those who do not report cannabis use. In terms of care, adults with CHS symptoms are likely to have numerous psychological and behavioral deficits that may affect their psychosocial functioning and make treatment engagement more challenging, thus requiring additional support beyond the treatment of their medical symptoms.

## Data Availability

The data will be despoited in the National Data Archive maintaind by the National Institutes of Health USA

## Acknowledgements

Research reported in this publication was supported by the National Institute of Mental Health awards R01MH137443 (Ilgen, Hicks) and RF1MH137443 (Ilgen, Hicks), K18 MH135466 (Hicks) of the National Institutes of Health, a 2025 Pilot award from the University of Michigan Institute for Firearm Injury, and Research Career Scientist award RCS 19-333 (Ilgen) of the Department of Veterans Affairs. The sponsors had no role in the design, collection, management, analysis, or interpretation of data, the writing of the manuscript, or submission for publication. Dr. Hicks and Dr. Ilgen had full access to all the data in the study and take responsibility for the integrity of the data and the accuracy of the data analysis. The content is solely the responsibility of the authors and does not necessarily represent the official views of the National Institutes of Health or the Department of Veterans Affairs.

## REFERENCES

Allen, J. H., de Moore, G. M., Heddle, R., & Twartz, J. C. (2004). Cannabinoid hyperemesis: cyclical hyperemesis in association with chronic cannabis abuse. Gut, 53(11), 1566–1570. 10.1136/gut.2003.036350

Angulo, M. I. (2024). Cannabinoid Hyperemesis Syndrome. Jama. 10.1001/jama.2024.9716

Caulkins, J. P. (2024). Changes in self-reported cannabis use in the United States from 1979 to 2022. Addiction, 119(9), 1648–1652. 10.1111/add.16519

Coelho, J., Montagni, I., Micoulaud-Franchi, J. A., Plancoulaine, S., & Tzourio, C. (2023). Study of the association between cannabis use and sleep disturbances in a large sample of University students. Psychiatry Res, 322, 115096. 10.1016/j.psychres.2023.115096

Copeland, W. E., Hill, S. N., & Shanahan, L. (2022). Adult Psychiatric, Substance, and Functional Outcomes of Different Definitions of Early Cannabis Use. J Am Acad Child Adolesc Psychiatry, 61(4), 533–543. 10.1016/j.jaac.2021.07.824

Cyders, M. A., Littlefield, A. K., Coffey, S., & Karyadi, K. A. (2014). Examination of a short English version of the UPPS-P Impulsive Behavior Scale. Addict Behav, 39(9), 1372–1376. 10.1016/j.addbeh.2014.02.013

Cyranowski, J. M., Zill, N., Bode, R., Butt, Z., Kelly, M. A., Pilkonis, P. A., Salsman, J. M., & Cella, D. (2013). Assessing social support, companionship, and distress: National Institute of Health (NIH) Toolbox Adult Social Relationship Scales. Health Psychol, 32(3), 293–301. 10.1037/a0028586

Dutwin, D. (2024). Response Rate Transparency in U.S. Probability Household Panels. N. a. t. U. o. Chicago. https://www.norc.org/content/dam/norc-org/amerispeak/response-rate-transparency-in-panels.pdf

ElSohly, M. A., Mehmedic, Z., Foster, S., Gon, C., Chandra, S., & Church, J. C. (2016). Changes in Cannabis Potency Over the Last 2 Decades (1995-2014): Analysis of Current Data in the United States. Biol Psychiatry, 79(7), 613–619. 10.1016/j.biopsych.2016.01.004

Garcia, A. M., & Rodríguez, S. F. (2025). Cannabis hyperemesis syndrome: an emerging clinical and public health challenge. Eur J Hosp Pharm, 32(4), 393. 10.1136/ejhpharm-2024-004369

Group, W. A. W. (2002). The Alcohol, Smoking and Substance Involvement Screening Test (ASSIST): development, reliability and feasibility. Addiction, 97(9), 1183–1194. 10.1046/j.1360-0443.2002.00185.x

Hall, W., & Lynskey, M. (2020). Assessing the public health impacts of legalizing recreational cannabis use: the US experience. World Psychiatry, 19(2), 179–186. 10.1002/wps.20735

Hamel, J., Jones, D. N., Dutton, D. G., & Graham-Kevan, N. (2015). The CAT: A Gender-Inclusive Measure of Controlling and Abusive Tactics. Violence Vict, 30(4), 547–580. 10.1891/0886-6708.Vv-d-13-00027

Hasin, D. S. (2018). US Epidemiology of Cannabis Use and Associated Problems. Neuropsychopharmacology, 43(1), 195–212. 10.1038/npp.2017.198

Hasler, W. L., Alshaarawy, O., & Venkatesan, T. (2025). Cannabis use patterns and association with hyperemesis: A comprehensive review. Neurogastroenterol Motil, 37(3), e14895. 10.1111/nmo.14895

Hoch, E., Volkow, N. D., Friemel, C. M., Lorenzetti, V., Freeman, T. P., & Hall, W. (2025). Cannabis, cannabinoids and health: a review of evidence on risks and medical benefits. Eur Arch Psychiatry Clin Neurosci, 275(2), 281–292. 10.1007/s00406-024-01880-2

Ilgen, M. A., Price, A. M., Goldman, P., Hicks, B.M. (2026). Prevalence and Correlates of Symptoms of Cannabinoid Hyperemesis Syndrome in the United States.

Iqbal, M. N., Levin, C. J., & Levin, F. R. (2019). Treatment for Substance Use Disorder With Co-Occurring Mental Illness. Focus (Am Psychiatr Publ), 17(2), 88–97. 10.1176/appi.focus.20180042

Jack, B., Susi, A., Reeves, P., & Nylund, C. M. (2025). Increasing trends of cannabinoid hyperemesis syndrome in youth: The grass is not always greener. J Pediatr Gastroenterol Nutr, 80(4), 638–643. 10.1002/jpn3.12469

Jenkins, C. D., Stanton, B. A., Niemcryk, S. J., & Rose, R. M. (1988). A scale for the estimation of sleep problems in clinical research. J Clin Epidemiol, 41(4), 313–321. 10.1016/0895-4356(88)90138-2

Jiménez-Castillo, R. A., Arumugam, S., Remes-Troche, J. M., & Venkatesan, T. (2025). Cannabinoid hyperemesis syndrome: A review. Rev Gastroenterol Mex (Engl Ed), 90(2), 214–226. 10.1016/j.rgmxen.2025.02.002

Krebs, E. E., Lorenz, K. A., Bair, M. J., Damush, T. M., Wu, J., Sutherland, J. M., Asch, S. M., & Kroenke, K. (2009). Development and initial validation of the PEG, a three-item scale assessing pain intensity and interference. J Gen Intern Med, 24(6), 733–738. 10.1007/s11606-009-0981-1

Kroenke, K., Spitzer, R. L., & Williams, J. B. (2003). The Patient Health Questionnaire-2: validity of a two-item depression screener. Medical care, 41(11), 1284–1292. 10.1097/01.Mlr.0000093487.78664.3c

Kroenke, K., Spitzer, R. L., Williams, J. B., Monahan, P. O., & Löwe, B. (2007). Anxiety disorders in primary care: prevalence, impairment, comorbidity, and detection. Ann Intern Med, 146(5), 317–325. 10.7326/0003-4819-146-5-200703060-00004

Loganathan, P., Gajendran, M., & Goyal, H. (2024). A Comprehensive Review and Update on Cannabis Hyperemesis Syndrome. Pharmaceuticals (Basel), 17(11). 10.3390/ph17111549

Mannes, Z. L., Malte, C. A., Olfson, M., Wall, M. M., Keyes, K. M., Martins, S. S., Cerdá, M., Gradus, J. L., Saxon, A. J., Keyhani, S., Maynard, C., Livne, O., Fink, D. S., Gutkind, S., & Hasin, D. S. (2023). Increasing risk of cannabis use disorder among U.S. veterans with chronic pain: 2005-2019. Pain, 164(9), 2093–2103. 10.1097/j.pain.0000000000002920

Mattingly, D. T., Richardson, M. K., & Hart, J. L. (2024). Prevalence of and trends in current cannabis use among US youth and adults, 2013-2022. Drug Alcohol Depend Rep, 12, 100253. 10.1016/j.dadr.2024.100253

Meltzer, A. C., Morrison, C., Loganathan, A., Shahamatdar, S., Moon, A., Heidish, R., Makutonin, M., Ma, Y., Li, R., & Cooper, Z. D. (2025). Cannabinoid Hyperemesis Syndrome Is Associated With High Disease Burden: An Internet-Based Survey. Ann Emerg Med, 85(6), 521–525. 10.1016/j.annemergmed.2025.01.008

Miki, A., Tandon, M., Murad, D., Loughman, J., Gilman, J., & Jangi, S. (2025). Increasing Prevalence of Cannabinoid Hyperemesis Syndrome in Young Adults and Minority Populations. Am J Gastroenterol, 120(12), 2954–2956. 10.14309/ajg.0000000000003591

Murphy, B. A., Casto, K. V., Watts, A. L., Costello, T. H., Jolink, T. A., Verona, E., & Algoe, S. B. (2022). “Feeling Powerful” versus “Desiring Power”: A pervasive and problematic conflation in personality assessment? Journal of Research in Personality, 101, 104305. 10.1016/j.jrp.2022.104305

Rubio-Tapia, A., McCallum, R., & Camilleri, M. (2024). AGA Clinical Practice Update on Diagnosis and Management of Cannabinoid Hyperemesis Syndrome: Commentary. Gastroenterology, 166(5), 930–934.e931. 10.1053/j.gastro.2024.01.040

Salsman, J. M., Schalet, B. D., Park, C. L., George, L., Steger, M. F., Hahn, E. A., Snyder, M. A., & Cella, D. (2020). Assessing meaning & purpose in life: development and validation of an item bank and short forms for the NIH PROMIS(®). Qual Life Res, 29(8), 2299–2310. 10.1007/s11136-020-02489-3

Saunders, J. B., Aasland, O. G., Babor, T. F., de la Fuente, J. R., & Grant, M. (1993). Development of the Alcohol Use Disorders Identification Test (AUDIT): WHO Collaborative Project on Early Detection of Persons with Harmful Alcohol Consumption--II. Addiction, 88(6), 791–804. 10.1111/j.1360-0443.1993.tb02093.x

Simonetto, D. A., Oxentenko, A. S., Herman, M. L., & Szostek, J. H. (2012). Cannabinoid hyperemesis: a case series of 98 patients. Mayo Clin Proc, 87(2), 114–119. 10.1016/j.mayocp.2011.10.005

Solmi, M., De Toffol, M., Kim, J. Y., Choi, M. J., Stubbs, B., Thompson, T., Firth, J., Miola, A., Croatto, G., Baggio, F., Michelon, S., Ballan, L., Gerdle, B., Monaco, F., Simonato, P., Scocco, P., Ricca, V., Castellini, G., Fornaro, M., Murru, A., Vieta, E., Fusar-Poli, P., Barbui, C., Ioannidis, J. P. A., Carvalho, A. F., Radua, J., Correll, C. U., Cortese, S., Murray, R. M., Castle, D., Shin, J. I., & Dragioti, E. (2023). Balancing risks and benefits of cannabis use: umbrella review of meta-analyses of randomised controlled trials and observational studies. Bmj, 382, e072348. 10.1136/bmj-2022-072348

Sorensen, C. J., DeSanto, K., Borgelt, L., Phillips, K. T., & Monte, A. A. (2017). Cannabinoid Hyperemesis Syndrome: Diagnosis, Pathophysiology, and Treatment-a Systematic Review. J Med Toxicol, 13(1), 71–87. 10.1007/s13181-016-0595-z

Soto, C. J., & John, O. P. (2017). The next Big Five Inventory (BFI-2): Developing and assessing a hierarchical model with 15 facets to enhance bandwidth, fidelity, and predictive power. J Pers Soc Psychol, 113(1), 117–143. 10.1037/pspp0000096

Spindle, T. R., Bonn-Miller, M. O., & Vandrey, R. (2019). Changing landscape of cannabis: novel products, formulations, and methods of administration. Curr Opin Psychol, 30, 98–102. 10.1016/j.copsyc.2019.04.002

Stjepanović, D., Kirkman, J., & Hall, W. (2025). Rare but relevant: Cannabinoid hyperemesis syndrome. Addiction, 120(2), 380–384. 10.1111/add.16693

STRAUS, M. A., Hamby, S. L., Boney-McCoy, S., & Sugarman, D. B. (1996). The Revised Conflict Tactics Scales (CTS2):Development and Preliminary Psychometric Data. Journal of Family Issues, 17(3), 283–316. 10.1177/019251396017003001

Swartz, J. A., & Franceschini, D. (2025). Cannabinoid Hyperemesis Syndrome, 2016 to 2022. JAMA Netw Open, 8(11), e2545310. 10.1001/jamanetworkopen.2025.45310

Wang, Q., Qin, Z., Xing, X., Zhu, H., & Jia, Z. (2024). Prevalence of Cannabis Use around the World: A Systematic Review and Meta-Analysis, 2000-2024. China CDC Wkly, 6(25), 597–604. 10.46234/ccdcw2024.116

Ware, J., Jr., Kosinski, M., & Keller, S. D. (1996). A 12-Item Short-Form Health Survey: construction of scales and preliminary tests of reliability and validity. Medical care, 34(3), 220–233. 10.1097/00005650-199603000-00003

